# Statistical Modeling of deaths due to COVID-19 influenced by social isolation in Latin American countries

**DOI:** 10.1101/2021.04.05.21254941

**Authors:** Rafael André da Silva, Luiz Philipe de Souza Ferreira, Jean Michel Rocha Sampaio Leite, Fernanda Assunção Tiraboshi, Thiago Maciel Valente, Vinicius Moraes de Paiva Roda, Jeniffer Johana Duarte Sanchez

**Affiliations:** Institute of Biomedical Sciences, University of São Paulo (ICB/USP), São Paulo, SP, Brazil; Paulista School of Medicine, Federal University of São Paulo (EPM/UNIFESP), São Paulo, SP, Brazil; School of Public Health, University of São Paulo (FSP/USP), São Paulo, SP, Brazil; University of Fortaleza (UNIFOR), Fortaleza, CE, Brazil; Federal University of Ceará (UFC), Fortaleza, CE, Brazil

**Keywords:** COVID-19, SARS-CoV-2, Coronavirus, Latin American countries, Social isolation, Government stringency index

## Abstract

Social isolation is extremely important to minimize the effects of a pandemic. Latin American (LA) countries have similar socioeconomic characteristics and health system infrastructures. These countries face difficulties to deal with the COVID-19 pandemic and some of them had very high death rates. Government stringency index (GSI) of twelve LA countries was gathered from the Oxford COVID-19 Government Response Tracker (OxCGRT) project. GSI is calculated considering nine metrics of social distancing and isolation measures. Population data from the United Nations Population Fund and number of deaths data was collected from the dashboard of the World Health Organization (WHO). We performed an analysis of the period March-December using a mixed linear model approach. Peru, Brazil, Chile, Bolivia, Colombia, Argentina and Ecuador had the highest death rates with an increasing trend over time, while Suriname, Venezuela, Uruguay, Paraguay and Guyana had the lowest ones, which remained steady. GSI in most countries followed the same pattern during the analyzed months. i.e., high indices at the beginning of the pandemic and lower ones in the last evaluated months, while the number of deaths increased over the whole period. Almost no country kept its GSI high for a long time, especially from October to December. Time and GSI as well as their interaction were highly significant. As their interaction increases, death rate decreases. In conclusion, a higher GSI at the start of the pandemic COVID-19 would impact on the decrease in the number of deaths over time, in LA countries.

## Introduction

The COVID-19 pandemic has affected healthcare systems and caused collapses across the globe. In Latin America (LA), the first case of SARS-CoV-2 infection was recorded on February 25th in the City of São Paulo. In less than a month after the first case, all LA countries had confirmed cases of COVID-19 ^1,2^.

The LA region has several obstacles that make it difficult for countries to take action against the spread of the virus. Precarious conditions, such as poverty, lack of hospital infrastructure, low sanitary conditions, high prevalence of chronic diseases and government’s tardy responses are factors that make it difficult to prevent contamination by the virus, **s**o that they facilitate transmission and directly impact the hospital system^3–5^. Through predictive models’ studies, it has been suggested that the virus could spread aggressively through LA^6,7^. Moreover, analyses of the initial cases of the COVID-19 pandemic in LA estimated an unfavorable scenario for the countries, and also evidenced aggressive dynamics of the disease outbreak in Brazil and Ecuador compared to Italy and Spain^7^. Above all, among the LA countries, Brazil was considered a major epicenter of the disease^8^.

Although there are measures aimed at reducing the spread of the new coronavirus such as social distancing, school closures, cancellation of public events and, sometimes, severe methods such as lockdown, these measures have been relaxed, in addition to noncompliance by the population and poor governmental management^9^. However, several current studies have already associated non-pharmaceutical interventions and lockdowns in particular to mortality, especially when measures are adopted early^10,11^. Although recent studies have estimated that non-pharmaceutical interventions are related to the number of deaths or the rate of infection of COVID-19 in different countries^12^, LA countries have not been the focus of these studies. LA countries should be analyzed with caution taking into account that most of the countries in America had difficulties in facing the pandemic^1,2,13^. In addition, Brazil is among the countries with the highest number of deaths caused by the virus^14^.

Considering that social distancing and isolation are important protective measures for the containment of SARS-CoV-2 infection, and that there is lack of studies discussing on demonstrating the relationship between the social isolation and death rate due to COVID-19 in LA countries, based on the Government Stringency Index (GSI) from the Coronavirus Government Response Tracker (OxCGRT)^15^, the objective of this work was to analyze the relationship between GSI and time, as well as death rate from COVID-19 in 12 LA countries, using a mixed linear model approach, especially looking at the measures adopted in the first year of the pandemic. Through this model, the need for maintaining social distancing and isolation measures over time at the start of the pandemic is explained and substantiated.

## Material and Methods

### Data Sources

Government stringency index (GSI) of twelve LA countries was developed by the Oxford University and gathered from the platform Policy Responses to the Coronavirus Pandemic on the website (https://ourworldindata.org/policy-responses-covid). This index represents the strictness of government policies and was calculated considering nine metrics of social distancing and isolation, such as school and work closures, stay-at-home requirements, transport restrictions, constraints on public gatherings, cancellation of public events, public information campaigns; restrictions on internal movements; and international travel controls. The index on any given day is calculated as the mean score of nine policy measures, each taking a value between 0 and 100. Hence, a higher GSI indicates a stricter response to the pandemic. The detailed methodology for calculating indices is described elsewhere (https://github.com/OxCGRT/covid-policy-tracker/blob/master/documentation/index_methodology.md). To use that data, we first calculated the mean of the GSI for each month. The number of cumulative monthly deaths data was collected from the dashboard of the World Health Organization (WHO). Population data were obtained from the United Nations Population Fund available on (https://www.unfpa.org/about-us) and used to calculate the proportion of the number of deaths for each country. This data was collected from the first day of March until the last day of December of 2020.

### Statistical analysis

We evaluated the relationship between GSI and time, and the death rates from COVID-19. Since we analyzed repeated measures over a period of time (time was uniformly measured across all countries), this is a longitudinal analysis requiring a mixed linear model approach,

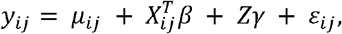

where

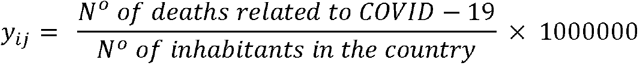

*μ* = mean of the death ratio adjusted by total population for each million inhabitants, *X*_*ij*_ = vector of covariates, *β* = vector of the regression parameters for the covariates. *Z* = matrix of covariates, *γ* = vector of random effects and *ε*_*i,j*_ = vector of random errors. *γ* and *ε* are uncorrelated.

In the model, the variables time (month) and GSI were considered both as fixed. In addition, the country was included as a random effect. All statistical analysis was performed under the most commonly used significance levels (1%, 5% and 10%) using the RStudio statistical software v.3.6.

## Results

We analyzed data of deaths related to COVID-19 in twelve LA countries in order to evaluate the relationship between death rates, GSI and time progression. In this context, time and GSI are useful to explain the dispersion of the data.

Figure 1a displays death rates from March to December. Peru, Brazil, Chile, Bolivia, Colombia, Argentina and Ecuador have high death rates, with an increasing trend over time, while Suriname, Venezuela, Uruguay, Paraguay and Guyana presented low death rates, which remained stable. Figure 1b displays GSI from March to December. It is noticeable that there was much fluctuation in the GSI for most countries, but with a large decrease from October to December. The only country whose GSI was maintained high for the whole period was Venezuela.

**Figure 1.**
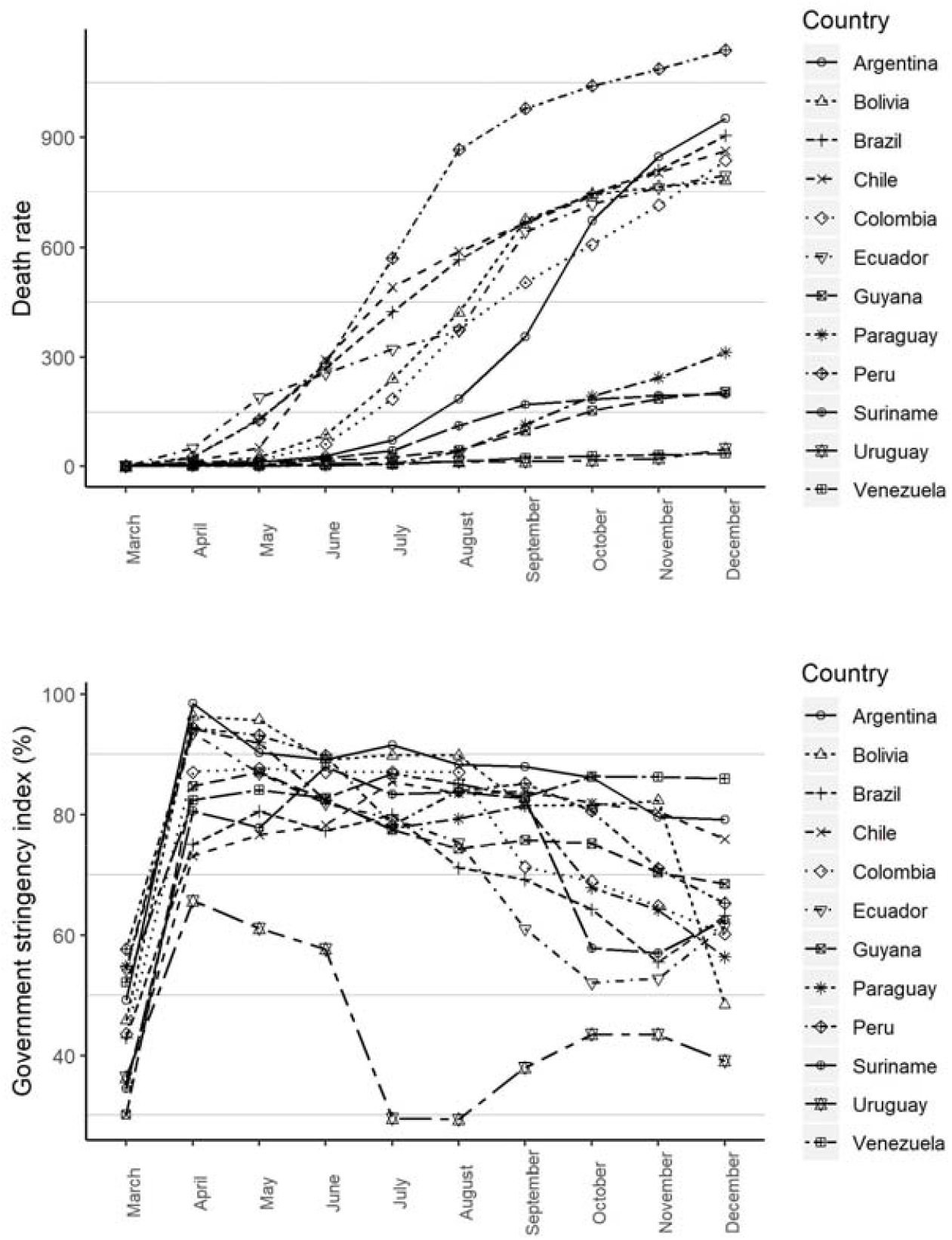
Death rates from COVID-19 and Government Stringency index from March to December 2020 in Latin American countries. Figure 1a: Death rates adjusted by population size for each country per million inhabitants. Figure 1b: Mean Government Stringency index reported as percentage.

Also, in Figure 1a, it is possible to observe the asymmetry of the data, so a skew-*t* distribution was adopted for the model’s error. Since the model presents a variable dispersion, we used a linear regression model for the dispersion.

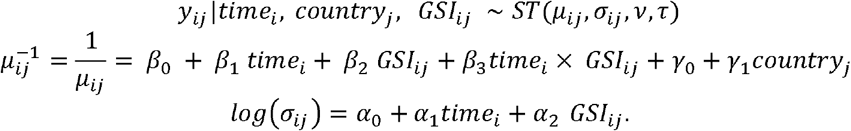

where *i* = 1, …, 10 represents each of the months, starting from March, and *j*= 1,…, 12 corresponds to each of the countries. *time*_*i*_ represents the *i*-th month, *country*_*j*_ represents the *j*-th country, *GSI*_*ij*_ represents the GSI in the *i*-th month in the *j*-th country. *σ*_*ij*_ corresponds to the standard deviation in the *i*-th month in the *j*-th country, with its corresponding parameters α. The random effects *γ*_0_ e *γ*_1_ have a normal distribution, and the response variable has a skew-*t* distribution with parameters, *γ*_*ij*_, *σ*_*ij*_,*v* and *τ*.

Table 1 displays the coefficient estimates of the mixed linear model and the dispersion model. The random effects are random variables that do not assume a single value, which means that its values cannot be displayed. Time and GSI, as well as their interaction, were highly significant to explain the death rate under all assumed significance levels. As the interaction of time and GSI increases, the death rate decreases.

**Table 1.** Estimates of the dispersion model and mixed linear model for death rates from COVID-19 in 2020 in Latin American countries.

|  | Estimate | Std Error | *t* Value | *p*-value |
| --- | --- | --- | --- | --- |
| $\mu$ | | | | |
| Intercept ($\beta_0$) | 491,633.0 | 69,250.3 | 7.099 | <0.005*** |
| Time ($\beta_1$) | -70,118.5 | 12,794.0 | -5.81 | <0.005*** |
| GSI($\beta_2$) | -6,721.4 | 984.6 | -6.826 | <0.005*** |
| Time×GSI ($\beta_3$) | 953.6 | 189.2 | 5.041 | <0.005*** |
| $\sigma$ | | | | |
| Intercept ($\alpha_0$) | -3.89 | 0.59 | -6.63 | <0.005*** |
| Time($\alpha_1$) | 0.72 | 0.04 | 16.67 | <0.005*** |
| GSI ($\alpha_2$) | 0.05 | 0.01 | 8.00 | <0.005*** |

Legend: GSI - Government stringency index. Significance levels: “\*\*\*” 0.001, “\*\*” 0.01, “\*” 0.05, “.” 0.1. The relationship between the predictors and the original response variable is inversely proportional i.e. a negative sign indicates an increase of death rates while the positive one indicates a decrease. Importantly, considering the interaction effect was significant, the main effects cannot be interpreted individually. For instance, the interpretation from the estimate obtained for the parameter associated with the interaction (953.6) is that for a given fixed month, if the GSI increases, the death rate decreases.

In Supplemental Figure S1a, the QQ-plot envelope shows there is no evidence that the skew-t distribution is inappropriate to explain the death rate for each million inhabitants. Other aspects of the model were analyzed by the quantile residuals (Supplemental Figure S2b), such as the correct specification of the model’s dispersion and distribution. We can conclude from these graphs that the model satisfies the assumptions so that the model specification is appropriate.

## Discussion

Robust evidence shows that, under most conditions, early adoption of stringent government non-pharmaceutical interventions is associated with a reduction in transmission and death^10–12^. Continued intervention should be considered to keep transmission of disease under control^10^. In addition it was estimated that GSI was able to decrease the number of deaths at different waves during the course of SARS-CoV-2 disease in different countries^12^. However, addressing the influence of this factor on death rates remains a big challenge because countries publish their testing data at different time points: some provide daily updates, while others provide only on a weekly-basis, and some only publish figures on an ad-hoc basis at longer intervals.

Based on GSI data extracted from the OxCGRT project ^15^, it is possible to propose statistical models to evaluate how closely these variables are related to time. Herein, the model shows that the relationship between time and GSI is highly significant. When analyzing time and GSI together, it was observed that, as the interaction of these two variables increases, a drop in the death rate is detected. For instance, according to this model, with a GSI set to 0 in March and a GSI set to 80 in April. i.e. a 80% increase in the strictness of government policies, we could have observed a reduction of approximately 32 deaths per million inhabitants. In Brazil, whose population is near 212 million, that would represent 6,784 lives that could have been saved at the beginning of the pandemic. This figure would be even higher in other months within the analyzed period. Furthermore, once these two variables were analyzed separately, as time increases, the rate of deaths per million inhabitants increases as well. Surprisingly, the same happens with the GSI reported by the countries. In light of these observations, we can make two hypotheses: 1) GSI alone may not entirely represent the reality regarding social isolation and the death rate from COVID-19, since this condition depends on other factors, such as the infrastructure of the countries’ public hospitals, government management, and the population’s compliance with the rules. 2) Restriction policies as measured by GSI do not have immediate effects and must be maintained over longer periods in order to decrease death rates by COVID-19. Hence, the problem is complex and deserves to be studied in detail taking into account other aspects that may be influencing the death rates.

LA countries present problematic issues, such as social inequality and less access to healthcare. However, complying with social isolation is difficult for individuals when work is the only source of income^13^. In our analyses, countries such as Peru, Brazil, Chile, Bolivia, Colombia, Argentina and Ecuador have the highest death rates. Peru presents low conditions to face a pandemic and even in lockdown at the beginning of the pandemic^2^, it presented high death rates. In a prediction study with data from the first 10 days of the pandemic, it was estimated that Peru had the lowest effective reproductive number (R_*t*_), parameter used to keep track of epidemics^7^, therefore, the country had these numbers accentuated during the pandemic period.

Brazil was the first LA country to report cases of COVID-19^16-18^. Since it presents favorable conditions to face a pandemic when compared to other LA countries, it was expected to have lower rates. However, in the present study, Brazil and Chile presented higher death rates, followed by Peru. It is important to consider that, although Peru’s president has played a relevant role helping to control the number of deaths from COVID-19, there has not been neither a national strategic plan to guide communication and educational health policies nor a large-scale awareness campaign to stimulate people to protect their health and abide to the protective measures. This lack of policies is also a current problem in Brazil^2^. For instance, through GSI e and COVID-19 Community Mobility Reports from Google, daily new cases and real time R_*t*_ were calculated, showing that Brazil is not doing very well regarding its response to COVID-19 pandemic^19^. Although Brazil presents a robust public health system and reasonable GSI, the high death rates may be deeply connected to inadequate policy management that has received several criticisms^5,13^. In comparison to Brazil, Suriname had a similar GSI but a low death rate, remaining stable over time. Except for Venezuela, no other country kept a high GSI for longer periods. Particularly, GSI decreased at the end of the year (October-December) in most countries, while death rates increased. On the other hand, isolation in Venezuela was maintained even in December (an atypical month because of Christmas and holiday season), and its death rates were low and remained unchanged over time.

According to the present analysis, Uruguay followed a relatively lower GSI than other LA countries but showed low death rates. Uruguay was a country that acted quickly, closing its borders and schools, with insertion of screening tests, reducing SARS-CoV-2 infections and controlling the outbreak very efficiently^20^. In contrast, Ecuador started with high social isolation, but a decrease in the isolation rate was observed later. On the other hand, Ecuador had a high mortality rate, which is accentuated over time even with the adoption of lockdown. In addition, it should be noted that this country had poor conditions of public health infrastructure at the beginning of the pandemic^2^. At the beginning of the COVID-19 pandemic, it had been already suggested that closing public transportation, workplaces and schools is particularly effective in reducing COVID-19 transmission^21^.

The rapidly evolving pandemic in LA countries is worthy of especial attention, considering their often weak and low stringency responses to the current sanitary crisis. In this study, GSI varies considerably in all LA countries over time. This variation can partially explain why these countries have been differently impacted by COVID-19. In spite of not specifically addressing and discussing the government policies adopted by each country, in this investigation, we successfully show that social distancing and isolation measured by GSI influences death rates from COVID-19 over time. For instance, the interaction between GSI and time can decrease the number of deaths, which demonstrates the importance of maintaining social distancing and isolation measures for longer periods, as opposed to what most LA countries did. Almost no country kept its GSI high for much time, especially from October to December. We did not expect to find different results since several studies support the idea that stringency policies are extremely important to contain the contagion or death of COVID-19^10–12^. Therefore, this is the first paper that discusses the importance of non-pharmaceutical interventions based on increased GSI could have directly impacted on the number of deaths in LA countries.

Our results have significant implications; however, some limiting aspects must be considered. 1) The GSI was extracted from the OxCGRT project. The curators of this database emphasized how challenging the collection of information on the exact data was due to the nature and extent of the policies of the different governments. This complex data set can obscure the qualitative differences in each of the nine metrics GSI measures across countries. In addition, many local and cultural factors can affect the implementation of interventions. 2) Our data provide a general interpretation of the influence of time and GSI on death rates in LA. Therefore, future studies can deepen the search for more specific interpretations for each country, taking into account local aspects and other metrics not covered here. 3) The numbers of deaths from COVID-19 can be easily underreported^18,19^, this is due to limited testing, problems in determining the cause of death and the way in which COVID-19 deaths are recorded. Hence, we cannot define the real impact of the GSI on death rates with perfect precision. 4) We know that the differences in population size between countries are often large, and the COVID-19 death count in more populous countries tends to be greater. Thus, in order to perform a more truthful comparison, we used the cumulative death data and calculated the death rate adjusted by the population of each country.

## Conclusions

We conclude that, in combination, time and GSI have beneficial effects on the decrease of death rates from COVID-19 in LA countries. Higher strictness of social distancing and isolation, as measured by the GSI, at the start of the pandemic, could have flattened mortality curves from COVID-19 over time, particularly from March to December 2020. Our statistical model explains and substantiates the need for maintaining social distancing and isolation measures over time during the pandemic.

## Supporting information

Supplementary Materials: Analysis of the model fit

## Data Availability

Data Availability Statement
Data openly available in a public repository that does not issue DOIs.
The data that support the findings of this study are openly available in:
1. Our World in Data [https://ourworldindata.org/policy-responses-covid]
2. Oxford COVID-19 Government Response Tracker (OxCGRT) [https://github.com/OxCGRT/covid-policy-tracker; https://covidtracker.bsg.ox.ac.uk/]
3. Dashboard of the World Health Organization (WHO) [https://www.who.int/]
References
1. Hasell J, Mathieu E, Beltekian D, Macdonald B, Giattino C, Ortiz-Ospina E, Roser M, Ritchie H. A cross-country database of COVID-19 testing. Sci Data. 2020 Oct 8;7(1):345. doi: 10.1038/s41597-020-00688-8.
2. Hale T, Angrist N, Goldszmidt R, Kira B, Petherick A, Phillips T, Webster S, Cameron-Blake E, Hallas L, Majumdar S, Tatlow H. A global panel database of pandemic policies (Oxford COVID-19 Government Response Tracker). Nat Hum Behav. 2021 Mar 8. doi: 10.1038/s41562-021-01079-8.
3. World Health Organization. WHO. Available from: https://www.who.int/

https://ourworldindata.org/policy-responses-covid

https://github.com/OxCGRT/covid-policy-tracker

https://covidtracker.bsg.ox.ac.uk/

https://www.who.int/

## Acknowledgments

Publication charges for this article were waived due to the ongoing COVID-19 pandemic.

## Authors’ addresses

Rafael André da Silva and Vinicius Moraes de Paiva Roda, Institute of Biomedical Sciences, University of São Paulo (ICB/USP), São Paulo, SP, Brazil. Luiz Philipe de Souza Ferreira, Paulista School of Medicine, Federal University of São Paulo, São Paulo, SP, Brazil. Thiago Maciel Valente and Fernanda Assunção Tiraboschi, University of Fortaleza (UNIFOR), Fortaleza, CE, Brazil. Jean Michel Rocha Sampaio Leite, School of Public Health, University of São Paulo (FSP/USP), São Paulo, SP, Brazil. Jeniffer Johana Duarte Sanchez, Federal University of Ceará (UFC), Fortaleza, CE, Brazil.

## Authors’ contributions

Rafael André da Silva: Conceptualization, Methodology, Data curation, interpretation, Writing - original draft, Supervision, critical review, Project administration. Luiz Philipe de Souza Ferreira: Writing - original draft, Methodology, Design of the study, Manuscript drafting, review, editing and Project administration. Jean Michel Rocha Sampaio Leite: Methodology, Data curation, Critical review, Interpretation, Writing – review, editing and Project administration. Fernanda Assunção Tiraboschi: Writing - review & editing. Thiago Maciel Valente: Data curation, Writing - original draft. Vinicius Moraes de Paiva Roda: Writing - original draft, critical review. Jeniffer Johana Duarte Sanchez: Conceptualization, Methodology, Formal analysis, Data curation, interpretation, Validation, Supervision and Project Head.

## References

1. Rodriguez-Morales AJ, Gallego V, Escalera-Antezana JP, et al. COVID-19 in Latin America: The implications of the first confirmed case in Brazil. Travel Med Infect Dis. 2020;35. doi:10.1016/j.tmaid.2020.101613

2. Garcia PJ, Alarcón A, Bayer A, et al. COVID-19 Response in Latin America. Am J Trop Med Hyg. 2020;103(5):1765–1772. doi:10.4269/ajtmh.20-0765

3. Miller MJ, Loaiza JR, Takyar A, Gilman RH. Covid-19 in latin america: Novel transmission dynamics for a global pandemic? PLoS Negl Trop Dis. 2020;14(5):3–7. doi:10.1371/JOURNAL.PNTD.0008265

4. Pablos-Méndez A, Vega J, Aranguren FP, Tabish H, Raviglione MC. Covid-19 in Latin America. BMJ. Published online 2020:369-370. doi:10.1136/bmj.m2939

5. Burky T. COVID-19 in Latin America Several. Lancet. 2020;(January):547.

6. Pacheco-Barrios K, Cardenas-Rojas A, Giannoni-Luza S, Fregni F. COVID-19 pandemic and Farr’s law: A global comparison and prediction of outbreak acceleration and deceleration rates. PLoS One. 2020;15(9 September):1–25. doi:10.1371/journal.pone.0239175

7. Caicedo-ochoa Y, Rebellón-sánchez DE, Peñaloza-rallón M. Effective Reproductive Number estimation for initial stage of COVID-19 pandemic in Latin American Countries. Int J Infect Dis. 2020;95(January):316–318.

8. Bermudi PMM, Lorenz C, Aguiar BS de, Failla MA, Barrozo LV, Chiaravalloti-Neto F. Spatiotemporal ecological study of COVID-19 mortality in the city of São Paulo, Brazil: Shifting of the high mortality risk from areas with the best to those with the worst socio-economic conditions. Travel Med Infect Dis. 2021;39(October 2020):101945. doi:10.1016/j.tmaid.2020.101945

9. Chen YT, Yen YF, Yu SH, Su ECY. An examination on the transmission of covid-19 and the effect of response strategies: A comparative analysis. Int J Environ Res Public Health. 2020;17(16):1–14. doi:10.3390/ijerph17165687

10. Flaxman S, Mishra S, Gandy A, et al. Estimating the effects of non-pharmaceutical interventions on COVID-19 in Europe. Nature. 2020;584(7820):257–261. doi:10.1038/s41586-020-2405-7

11. Brauner JM, Mindermann S, Sharma M, et al. Inferring the effectiveness of government interventions against COVID-19. Science (80-). 2021;371(6531). doi:10.1126/science.abd9338

12. Hale T, Angrist N, Hale AJ, et al. Government responses and COVID-19 deaths: Global evidence across multiple pandemic waves. PLoS One. 2021;16(7 July):1–14. doi:10.1371/journal.pone.0253116

13. The Lancet. COVID-19 in Latin America: a humanitarian crisis. Lancet. 2020;396(10261):1463. doi:10.1016/S0140-6736(20)32328-X

14. Naveca FG, Nascimento V, de Souza VC, et al. COVID-19 in Amazonas, Brazil, was driven by the persistence of endemic lineages and P.1 emergence. Nat Med. 2021;27(7):1230–1238. doi:10.1038/s41591-021-01378-7

15. Hale T, Angrist N, Goldszmidt R, et al. A global panel database of pandemic policies (Oxford COVID-19 Government Response Tracker). Nat Hum Behav. Published online 2021. doi:10.1038/s41562-021-01079-8

16. Philipe L, Ferreira DS, Valente TM, Tiraboschi FA. Description of Covid-19 Cases in Brazil and Italy. SN Compr Clin Med. Published online 2020:19-22.

17. Valente TM, Ferreira LPS, Silva RAD, Leite JMRS, Tiraboschi FA BM. Brazil Covid-19D: Change of hospitalizations and deaths due to burn injuryD? Burns. Published online 2020:9-11. doi:10.1016/j.burns.2020.10.009

18. de Souza WM, Buss LF, Candido D da S, et al. Epidemiological and clinical characteristics of the COVID-19 epidemic in Brazil. Nat Hum Behav. 2020;4(8):856–865. doi:10.1038/s41562-020-0928-4

19. Dongshan zhu SRM,, Xikun Han, Santo K. Social distancing in Latin America during the COVID-19 pandemic: an analysis using the Stringency Index and Google Community Mobility Reports. Int Soc Travel Med. Published online 2020:1-25.

20. Elizondo V, Harkins GW, Mabvakure B, et al. SARS-CoV-2 genomic characterization and clinical manifestation of the COVID-19 outbreak in Uruguay. Emerg Microbes Infect. 2021;10(1):51–65. doi:10.1080/22221751.2020.1863747

21. Vannoni M, McKee M, Semenza JC, Bonell C, Stuckler D. Using volunteered geographic information to assess mobility in the early phases of the COVID-19 pandemic: A cross-city time series analysis of 41 cities in 22 countries from March 2nd to 26th 2020. Global Health. 2020;16(1):1–9. doi:10.1186/s12992-020-00598-9

