## Supplementary figures and images for "Statistical Modeling of deaths due to COVID-19 influenced by social isolation in Latin American countries"

### Supplementary Materials: Analysis of the model fit

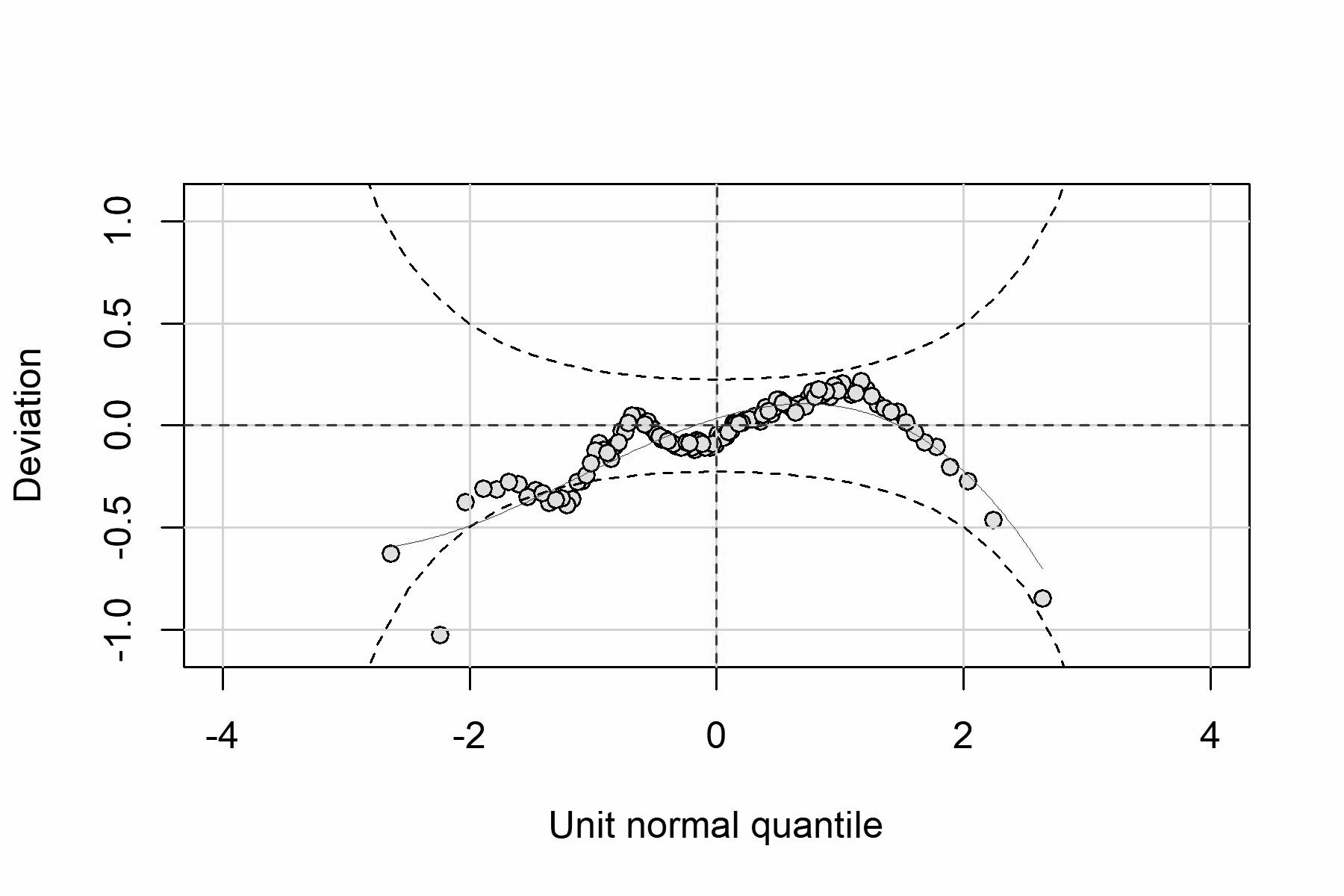

S1b)

S1a)

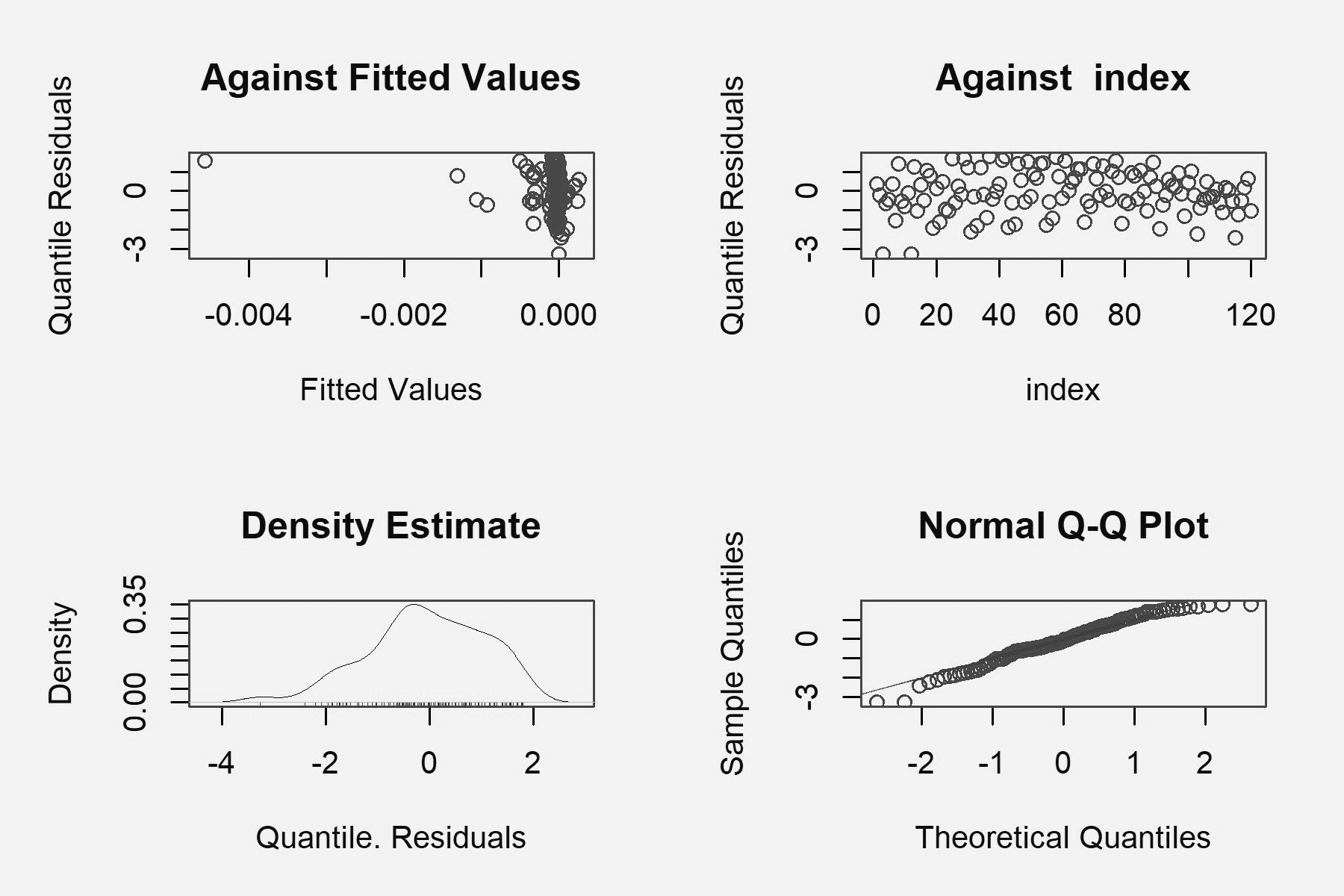
